# Modelling pooling strategies for SARS-CoV-2 testing in a university setting

**DOI:** 10.1101/2021.01.19.20248560

**Authors:** Gibran Hemani, Amy C Thomas, Josephine G. Walker, Adam Trickey, Emily Nixon, David Ellis, Rachel Kwiatkowska, Caroline Relton, Leon Danon, Hannah Christensen, Ellen Brooks-Pollock

## Abstract

Pre-symptomatic and asymptomatic transmission of SARS-CoV-2 are important elements in the Covid-19 pandemic, and until vaccines are made widely available there remains a reliance on testing to manage the spread of the disease, alongside non-pharmaceutical interventions such as measures to reduce close social interactions. In the UK, many universities opened for blended learning for the 2020-2021 academic year, with a mixture of face to face and online teaching. In this study we present a simulation framework to evaluate the effectiveness of different asymptomatic testing strategies within a university setting, across a range of transmission scenarios. We show that when positive cases are clustered by known social structures, such as student households, the pooling of samples by these social structures can substantially reduce the total cost of conducting RT-qPCR tests. We also note that routine recording of quantitative RT-qPCR results would facilitate future modelling studies.

## Introduction

In the midst of the COVID-19 pandemic, university students represent a demographic in the population who are likely to experience low rates of symptomatic infection^1^ while being in a high contact social setting^2,3^. Extensive testing is required to monitor and manage potentially high rates of SARS-CoV-2 infection within a student population, particularly to minimise transmission to vulnerable individuals and groups, both within and outside of the student population^4^. Recent modelling work demonstrated that very frequent testing would be required to impact transmission^5^, which comes at a high cost. The reverse transcriptase quantitative polymerase chain reaction (RT-qPCR) test has been in widespread use globally for the detection of viral RNA taken from saliva or nasal and throat swabs^6^. One strategy to reduce the financial cost of regular mass testing is to pool samples^7^ - a single test is performed on a group of individuals, which can optionally be converted into Dorfman’s algorithm in that if the group tests positive, follow up tests are performed on the individuals in the group to identify those infected^8^. Another benefit of this approach is that pooling reduces test reagent use, the supply of which may be outstripped by demand. More complex pooled testing algorithms have been proposed that could offer higher efficiencies but are difficult in practice to implement^9^.

There have been several studies evaluating pooling approaches for RT-qPCR for SARS-CoV-2 in order to identify cases that may be asymptomatic or pre-symptomatic^9,10^, and a pilot programme for pooled testing within universities is now underway^11^. In this analysis we explore the key decisions needed to design an effective test pooling strategy.

### Size of the pool

There exists a tension between reducing the cost by pooling samples versus the potential adverse impact of pooled samples on the sensitivity of the test. In the context of RT-qPCR, if only one infected individual is present amongst the samples in a pool, the samples from the uninfected individuals will dilute the viral load within, potentially increasing false negative rates. The trade off between cost and sensitivity when using Dorfman’s algorithm is strongly related to prevalence. When prevalence is low, few pools will contain infected samples, and so the number of pools that require follow-up tests will be low. Hence it will be beneficial to have pools with relatively large numbers of individuals in this scenario. By contrast, when prevalence is high, we might expect larger pool sizes to frequently test positive, and so the number of follow up tests required to identify specific individuals who are infected will be large. This dynamic can inform optimal pool sizes. A more complex follow up strategy that starts with large pools and iteratively creates sub pools to minimise testing numbers will have the disadvantage of a longer lead time between sample collection and test result, which has practical consequences because informing the infected individual swiftly is of importance in limiting spread^12^.

### Household or contact-based pooling

Another consideration is who is pooled with whom as it is likely advantageous for the test sensitivity to pool individuals who are close contacts and therefore liable to be all infected or all uninfected at the same time. In the university setting, a method for reducing social contacts is to assign students to small ‘living circles’ (e.g. five people per living circle). Because transmission often occurs in households, we might expect clustering of cases within living circles. Testing by living circle could improve test sensitivity under the assumption that if one person is infected then others are likely to be also, thus the dilution of the viral load is minimised. This pooling strategy could also reduce costs because fewer pools will be detected as positive than under random pooling, and therefore fewer pools will require follow up tests to identify infected individuals.

### Follow up testing

A third consideration is how follow up of positive pooled samples, to determine which individuals are specifically infected, impacts overall performance. In the UK this is currently mandated for positive pooled tests but deserves exploration. If tests are performed by living circle, then the follow up tests may be rendered largely moot if everyone within a living circle is expected to self-isolate when any one member of the living circle is tested positive. However, the impact on behaviour change if only returning pool-level test results to individuals needs to be quantified, such as whether individuals assume they have been infected and are immune.

Additional to pooling of samples in RT-qPCR assays, other low-cost screening methods are emerging. The lateral flow device operates by detecting viral antigens directly^13^, and though it has been reported to have lower sensitivity compared to RT-qPCR^14,15^ it may still be cost effective, and of sufficiently high overall sensitivity if the test is administered twice over three days^11^.

At the University of Bristol students within halls of residence are divided into living circles – groups within which individuals are permitted to mix as if in a household. Here we analyse the sensitivity and cost of pooling samples for SARS-CoV-2 testing by RT-qPCR within a university setting to represent a universal testing strategy that would identify asymptomatic infection. We use data on the sizes of living circles to simulate clustering of infections, allowing for varying levels of social contacts within and between living circles. We explore the effects of pool size and living-circle based pooling, follow up testing strategy, and heterogeneity of viral load between individuals.

## Methods

### Modelling overview

Our objective is to simulate performing RT-qPCR for presence of SARS-CoV-2 in samples from all individuals under different disease transmission scenarios. There are four main components to the model – viral load sampling, disease transmission between students, pooling allocation of collected samples, and testing performance. We compare different aspects of testing performance across three strategies – per-individual testing, pooled testing, or pooled testing with per-individual follow-up in positive pools.

### Data

The samples generated in these simulations are allocated into living circles and halls of residence that match the distributions from undergraduate students at the University of Bristol. In total there are 8,477 students allocated to 1,529 living circles, divided across 37 halls of residence. The median number of students per living circle is 5, and the maximum is 44. The distribution of students per living circle is shown in **Supplementary Figure 1**.

### Heterogeneity of viral load and RT-qPCR detection

To generate heterogeneity in the viral load of infected individuals, we first developed a model based on RT-qPCR mechanics whereby each individual *i* is assumed to have some quantification cycle value (*C*_*q,i*_) – defined as the number of cycles (*n*) required to reach the fluorescence threshold 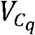 at which viral RNA is detected and considered positive within the RT-qPCR process. Quantification cycle value is a function of their sampled viral load *V*_*i,t*_ and the RT-qPCR efficiency (*E*), assuming each sample is tested individually. The efficiency can vary by sample and reflects, for example, random contaminants within the sample. In order to test positive, an individual’s *C*_*q,i*_ value must be below 35 (i.e. amplification of viral RNA reaches a detectable fluorescence level in the RT-qPCR reaction before the 35^th^ cycle). The exponential growth phase of the RT-qPCR kinetics can be modelled^16^ such that the number of viral particles after *n* cycles is given by

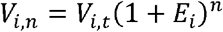

where *E*_*i*_ is the efficiency of the reaction for sample *i*. We can therefore express the number of cycles (*n* = *C*_*q,i*_) required to reach the viral fluorescence threshold 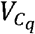 as

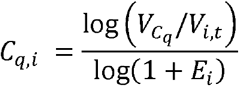

We generate a per-individual viral load time-course, where the viral load changes over time similar to that shown in Mina et al (2020). Specifically, we assume that each infected individual has a viral load at time point *t* that is

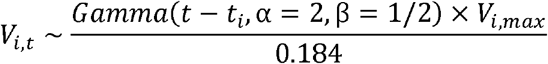

A value of *t*-*t*_*i*_, the number of days individual *i* has been infected for, that lies between 0 and 21 days^17^ is sampled uniformly to be the time point of the infection course the individual is tested. *V*_*i,max*_ represents the individual’s maximum viral load, and the denominator (0.184) is the maximum value of the gamma function with parameters α= 2 and β= 1/2. Therefore, the heterogeneity in viral load between individuals is generated by a) when during their infection course they are tested and b) the peak viral load over the course of the infection. Assuming a given individual-specific RT-qPCR efficiency (*E*_*i*_), the maximum *C*_*q*_ value per individual is

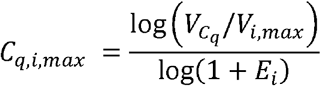

- In order to obtain realistic maximum viral load values, we generate *C*_*q,i,max*_ and *E*_*i*_ such that
- The distribution of *C*_*q*_ amongst positive cases resembles that from previous publications^18^
- The distribution of *E* values resembles that from previous publications^6,16,19^
- The maximum sensitivity (*S*) of undiluted samples (*D*= 1) matches those *s* (*D* = 1) = 0.98
- The maximum sensitivity of 10x diluted samples match those estimated from calibration tests of *s* (*D* = 10) = 0.80

The value of 98% maximum sensitivity for 1x dilution is taken from Arevalo-Rodriguez et al (2020)^20^ as the 90% upper bound of sensitivity across 34 studies reporting RT-qPCR sensitivity, and also supported by Visseaux et al (2020)^21^. The value of 80% sensitivity at 10x dilution is estimated on the assumption that the *C*_*q,i*_ value will increase by 3.32 and that 20% of the values at 1x dilution will be shifted to be above the threshold of 35 cycles (assuming 100% efficiency).

In order to determine the distributions for *C*_*q,max*_ and *E* we assume that both are independently beta distributed such that

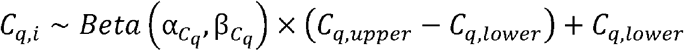

and

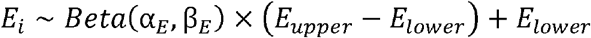

Following Kudo et al (2020)^19^ and Vogels et al (2020)^6^ we fix *E*_*upper*_ = 1.4 and *E*_*lower*_ = 0.9, but we must determine values of 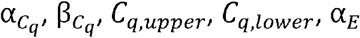 and β_*E*_ to satisfy the conditions stated above. To achieve this we use a general optimisation function which performs the following procedure

1. Sample values of *C*_*q,i,max*_ for 500,000 individuals (an arbitrarily large number to reduce sampling error) for a set of values for 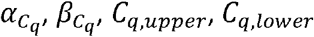
2. Sample values of *E*_*i*_ for a set of values for *α*_*E*_ and *β*_*E*_
3. Calculate the viral load per individual 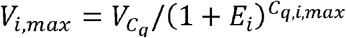 where the reference value 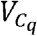 is set to 1 arbitrarily
4. Calculate the viral load in the sample after 10x dilution by dividing V_i,0_ (D= 10) = *V*_*i,max*_/10
5. Calculate the resultant expected 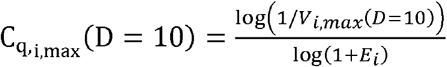
6. Calculate the loss function *L* = (*P* (*C*_*q,i,max*_ (*D* = 10) − *S* (*D* = 10))^2^ + (*P* (*C*_*qi,max*_ (*D*= 1) −*S* (*D*= 1)) ^2^
7. Repeat from 1 to minimise the value of *L*.

We use the default settings in the *optim* function in R 4.0.2^22^ to solve this optimisation function, resulting in the following parameter values:

- 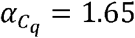
- 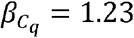
- *C*_*q,upper*_ = 35.5
- *C*_*q, lower*_ = 15.8
- *α*_*E*_ = 1.75
- *β*_*E*_ = 1.98

These values give rise to the distributions of viral load and RT-qPCR kinetics shown in **Supplementary Figure 2**. Reproducible code to determine these parameters are provided in https://explodecomputer.github.io/covid-uob-pooling/cq.html.

### Transmission model

To generate clustering of cases according to social structures amongst the students, such as living circles and halls of residence, we use a single time-step agent-based transmission model to capture transmission that can occur within a week – approximately the serial interval for COVID-19. At time step 0, a random set of individuals from the total population is selected to be infected according to some initial prevalence *P* (0). The individual reproduction number, or the number of people each individual goes on to infect is set as

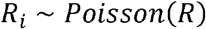

Where *R* is the reproduction number. We allow the *R* value to vary across simulation scenarios. The individuals who are infected are determined by contact patterns and the level of containment assumed for a particular simulation scenario. We assume that transmission can occur in one of two contexts: within the same living circle, or anywhere else. Hence, at time step 1, a new set of individuals will be infected, leading to an updated prevalence *P* (1). **Table 1** lays out the probability that a transmission event occurs in each context, based on different data sources or assumptions. For the base case analysis, we use the transmission event probabilities based on the CON-QUEST survey^23^, and present sensitivity analyses in **Supplementary figure 3**.

**Table 1:** Probabilities of transmission events for an infected individual to infect others in the same living circle or elsewhere. Each row represents a different containment scenario.

| **Containment scenario** | **Description** | **Same living circle (household)** | **Anywhere else** |
| --- | --- | --- | --- |
| **High containment** | Hypothetical circumstance where most transmission outside of living circles is limited | 0.9 | 0.1 |
| **Social contact survey (students)<sup>38</sup>** | Based on contact patterns from a pre-pandemic survey | 0.38 | 0.62 |
| **CON-QUEST survey<sup>2</sup>** | Based on student contact patterns surveyed during the pandemic (June to November 2020) | 0.76 | 0.24 |

### Testing pool allocation

We adopted two strategies to assign individuals to testing pools. First, we allowed assignment to be random. Second, we attempted to maximise grouping within test pools given living circles. The living circle sizes within the University of Bristol vary from 1 to 44, with a median of 5. However, within each simulation scenario the pool size is fixed (though we try different pool sizes between simulation scenarios). To maximise grouping of living circle members within a testing pool we used a simple bin packing algorithm, *binPack* from the *BBmisc* R package (version 1.11)^24^, which uses a greedy algorithm to maximise pool occupancy whilst minimising distribution of living circle members across multiple pools.

### Testing model

For all individuals in the sample the values of *E*_*i*_ and *C*_*q,i*_ are sampled as described in the ‘Viral Load’ section above, which gives rise to per individual viral loads *V*_*i,t*_. To calculate the test result for a pooled sample *j*, we estimate the pool viral load across samples *k* in a pool of size *D* samples using

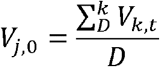

and estimate whether the pool would be detected as positive given the inequality *C*_*q,j*_ ≤ 35 where

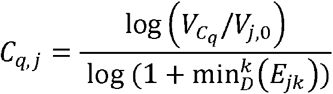

Note that we assume that the reaction efficiency of the pooled sample is equal to the lowest efficiency amongst the *k* samples contributing to the pool, in part to be conservative in evaluating the efficacy of pooling, and in part on the assumption that efficiency is adversely impacted by sample impurities.

We assume a diagnostic specificity of 99.5% for a single RT-qPCR test, and therefore the pool+followup results will have a false positive rate of 0.005^2^.

The cost of testing was estimated based on consumable costs alone, assuming that overhead or facility costs would be the same for any model of testing, and that each laboratory would have sufficient capacity and equipment (nucleic acid extraction and qPCR instruments, and storage freezers) to conduct the tests. We determined the cost of sample collection to be £3.47 (comprising of saliva sampling funnel (£1.86; Isohelix), collection tube (£1.42; Isohelix), storage tube, and pipette tip (£0.19)) and the cost of a PCR test to be £12.46 (based on 1 extraction using QIAsymphony® (QIAGEN) kits at £4.46 per sample, and one PCR reaction using the Altona SARS-CoV-2 RealStar RT-PCR Kit 1.0 at £8.00 per sample.

### Lateral flow device

For the lateral flow device tests (LFD) we assume specificity of 99.68% and a sensitivity that is a function of viral load, *S*(*C*_*q,i*_). The Joint PHE Porton Down & University of Oxford SARS-CoV-2 test development and validation cell^13^ provides a mapping of RT-qPCR kinetics against LFD sensitivity, which we interpolated using a sigmoidal model using the SSlogis function in R (**Supplementary figure 4**, https://explodecomputer.github.io/covid-uob-pooling/lfd.html). The probability of testing positive for infected individuals is sampled as

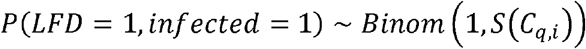

and for uninfected individuals

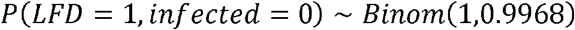

In practice the LFD protocol is to perform the test twice, across 3 days, and as such we also estimate the sensitivity of getting at least one positive result given changes in viral load across this time window. We assume a cost of £5 per LFD test^25^.

### Simulation setup

We explore the performance of pooling strategies under the following simulation scenarios

Initial prevalence: 0.001, 0.01, 0.05

R value: 0.8, 1, 3

Containment: ‘high’, ‘medium’, ‘low’

Pool sizes are chosen to be 2, 3, 4, 5, 10, 15, 20, 25 or 30, and individuals are allocated to pools according to their living circles or at random. This results in 486 simulation scenarios, each of which is repeated 100 times.

The simulations were all conducted using R 4.0.2 and made reproducible via a Snakemake pipeline^26^. Code can be accessed here: https://github.com/explodecomputer/covid-uob-pooling

## Results

### Pooled testing volume depends on epidemiological context

A major motivation for a pooled testing strategy is to reduce the financial cost by reducing the number of assays that need to be performed. In a simple pooling approach that does not follow-up positive pooled samples to determine the specific individuals infected, this reduction in costs will be related to the number of samples included in the pool, but results in all individuals in the pool behaving as if they are infected. However, as is well established, if positive pools are to be followed up then the total number of tests could drastically increase when prevalence is high (**Figure 1**), particularly when pool sizes are larger. For example, when initial prevalence is at 5% and the R value is high, a strategy of pool size of 20 comprising random samples will require as many tests as a simpler approach of testing everybody, if positive pools are followed up for individual testing. However, if we pool based on living circle in this context where most cases are clustered, then fewer pools will contain positive individuals, and the number of tests could be attenuated by 15%.

**Figure 1:**
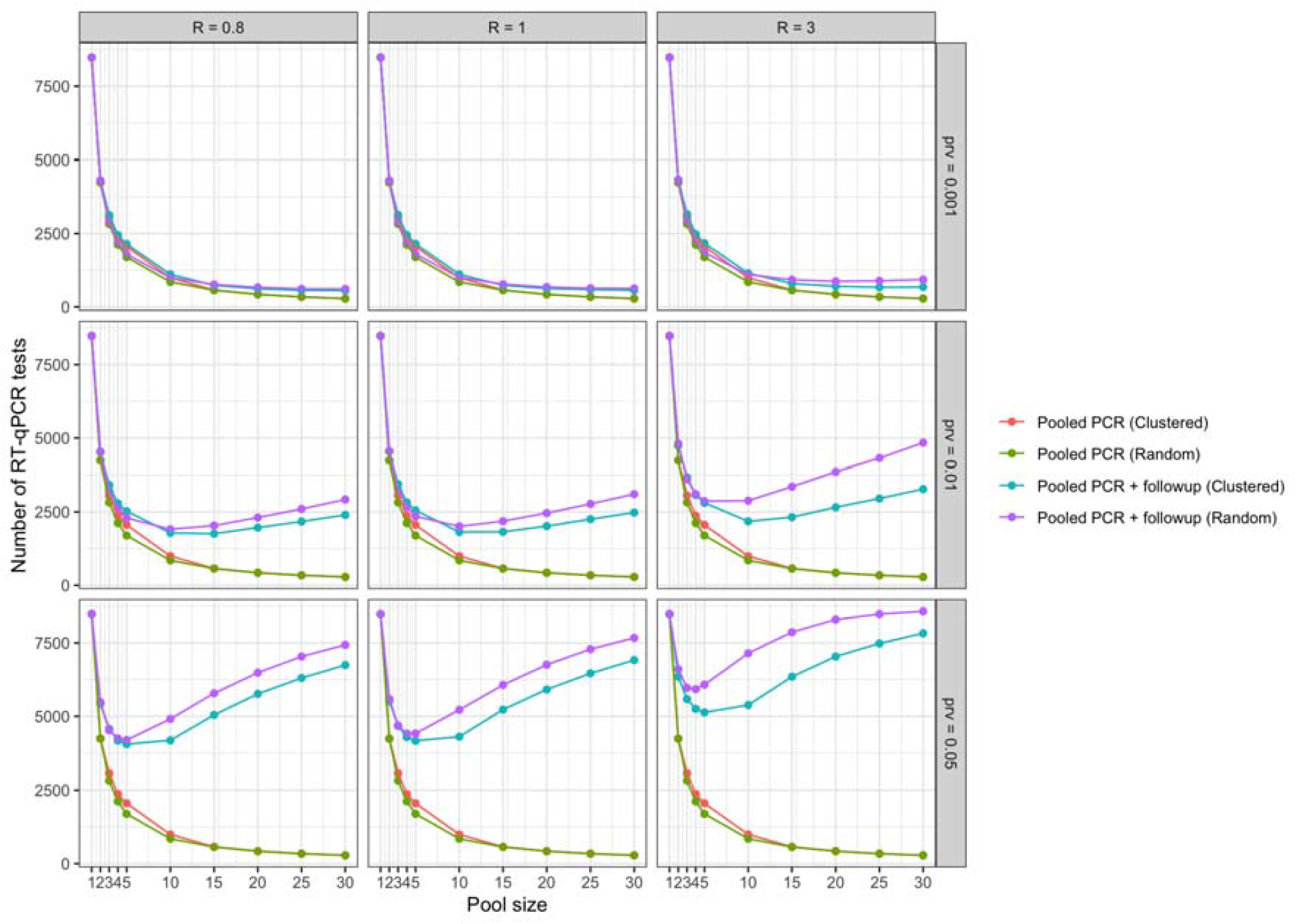
Comparison of number of tests required across different RT-qPCR strategies. The x-axis represents the pool size, with a value of 1 corresponding to a test per individual. The y-axis represents the number of tests to be performed. Rows of boxes represent the starting prevalence before transmission occurs, and columns of boxes represent the degree of transmission in terms of reproduction number (R).

### Test sensitivity

The rate at which infected individuals are tested positive varies across the various simulation scenarios. Pooling samples reduces test sensitivity as expected, but how the performance depends substantially on the loss is related to clustering of cases, pooling strategy and prevalence (**Figure 2**). Higher clustering of cases reduces loss of sensitivity as pool size increases, because the dilution of nucleotides is reduced when clustered cases are pooled together. A similar amelioration of the impact of dilution occurs as the prevalence increases. The improvement in sensitivity achieved by pooling by living circle is particularly pronounced when disease transmission is largely restricted to be within living circles, as this minimises isolated positive cases within pools. The sensitivity of LFD improves substantially when it is applied twice; however, in most scenarios pooling approaches will outperform the sensitivity of LFD.

**Figure 2:**
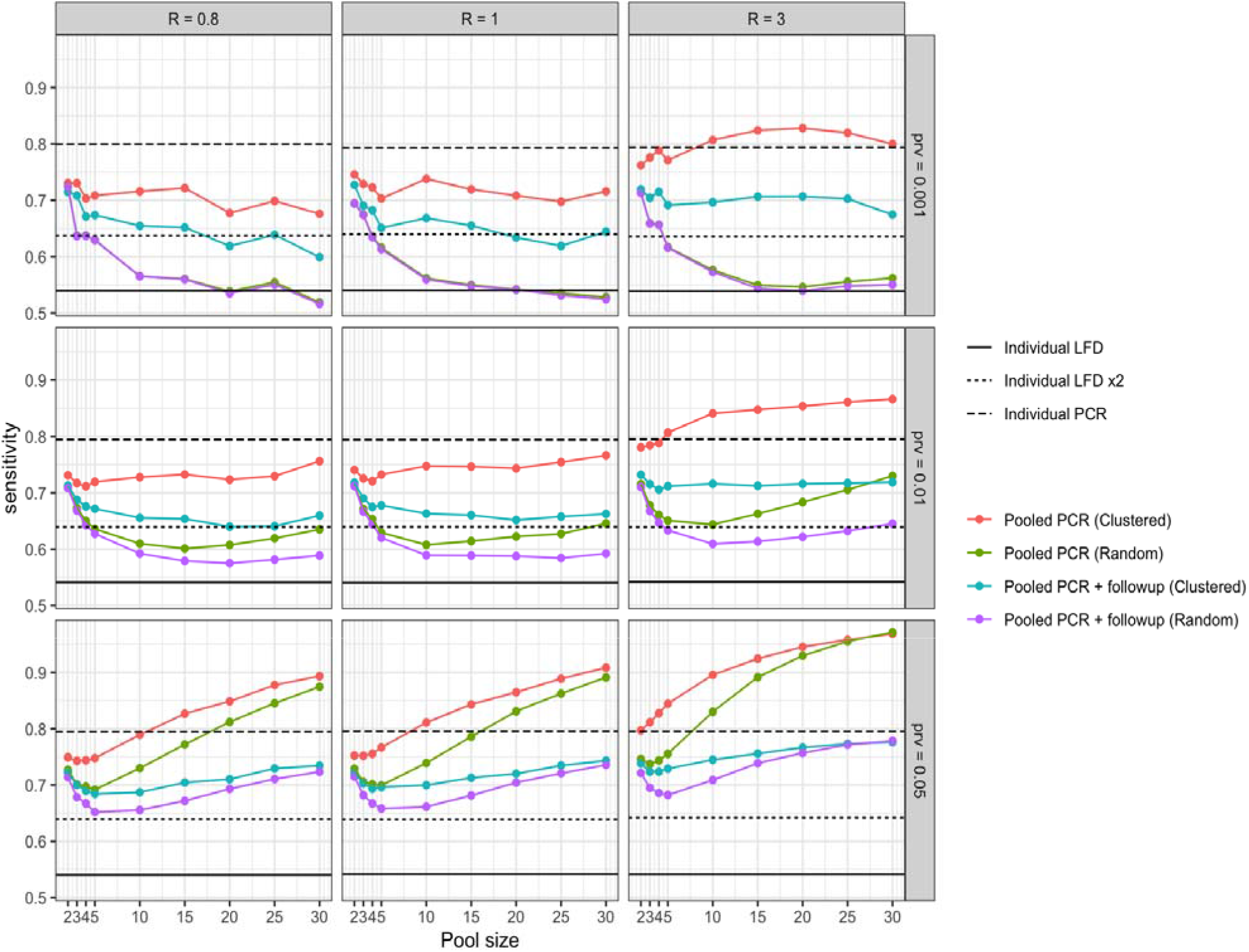
Comparison of test sensitivity across different testing strategies. The x-axis represents the pool size. Horizontal lines represent different individual-based testing approaches. The y-axis represents the sensitivity, defined as the proportion of infected individuals who are detected as positive. Rows of boxes represent the starting prevalence before transmission occurs, and columns of boxes represent the degree of transmission in terms of reproduction number (R).

### Cost effectiveness of testing strategies

We based the cost effectiveness of a particular strategy on its positive predictive value (PPV), defined as the probability, given a positive test result, that an individual really has the disease. PPV is a function of prevalence, sensitivity and specificity. **Figure 3** shows that the strategy that maximises the PPV to cost ratio varies substantially by prevalence. When prevalence is low then pooled tests are likely to reduce the proportion of false positives and therefore can improve on individual based tests. When prevalence is high, the sensitivity is relatively similar across all strategies, the false positive rate is a small fraction of total positives, and so the low cost of LFD appears favourable.

**Figure 3:**
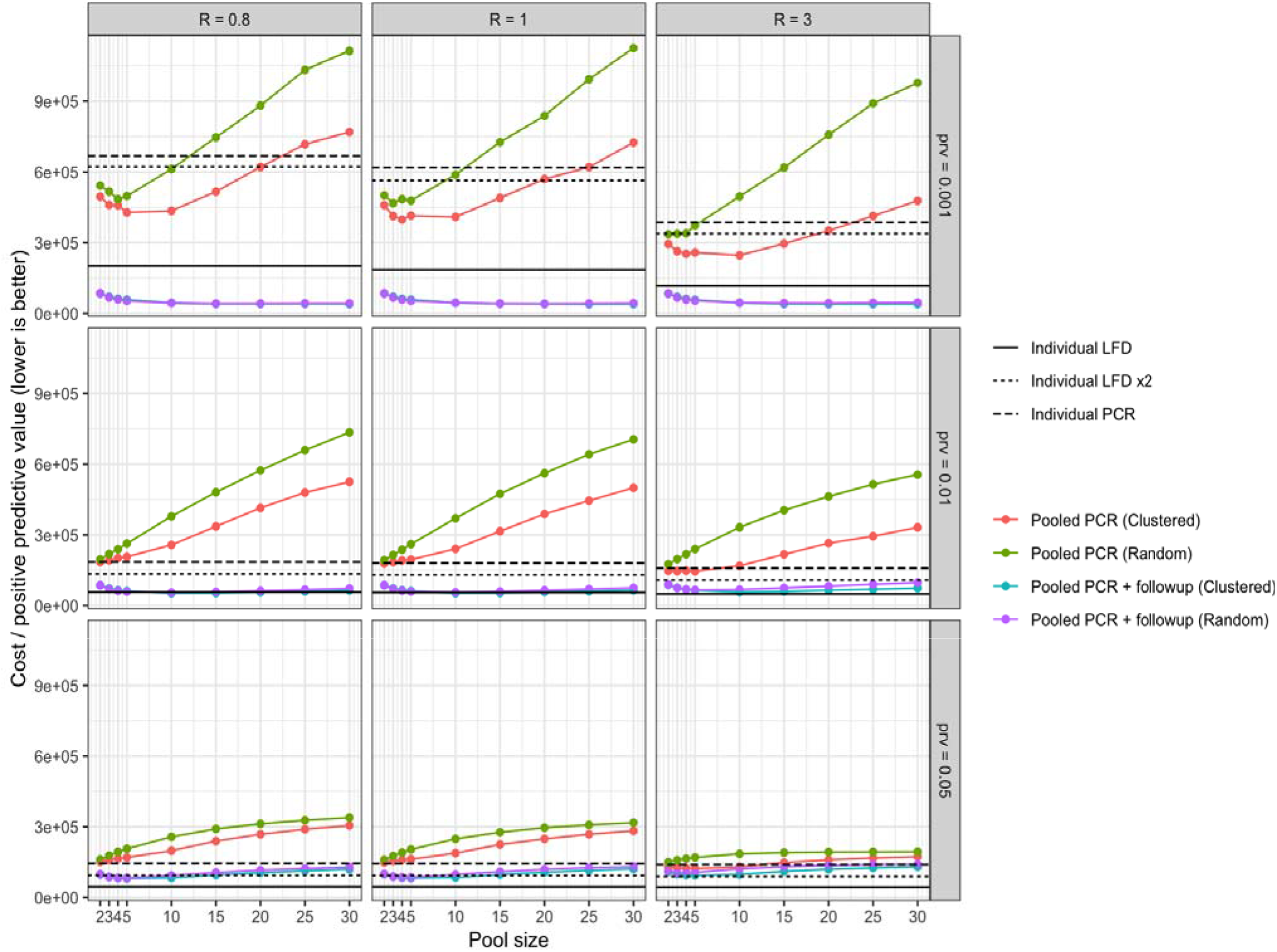
Comparison of cost per test effectiveness (in terms of positive predictive value) across different testing strategies. The x-axis represents the pool size. Horizontal lines represent different individual-based testing approaches. The y-axis represents the cost per positive predictive value (PPV) achieved. Rows of boxes represent the starting prevalence before transmission occurs, and columns of boxes represent the degree of transmission in terms of reproduction number (R).

### Estimating prevalence

Large scale testing can also be valuable as a surveillance tool. We evaluated the accuracy prevalence estimates for different testing strategies (**Figure 4**) and found that individual tests and pooled tests with follow ups offered good estimates of the prevalence. However, if pooled tests without follow up were to be used to estimate the prevalence then assumptions would need to be made about the proportion of samples within a test that are likely to be positive in order for that to be a cost-effective way of estimating prevalence accurately.

**Figure 4:**
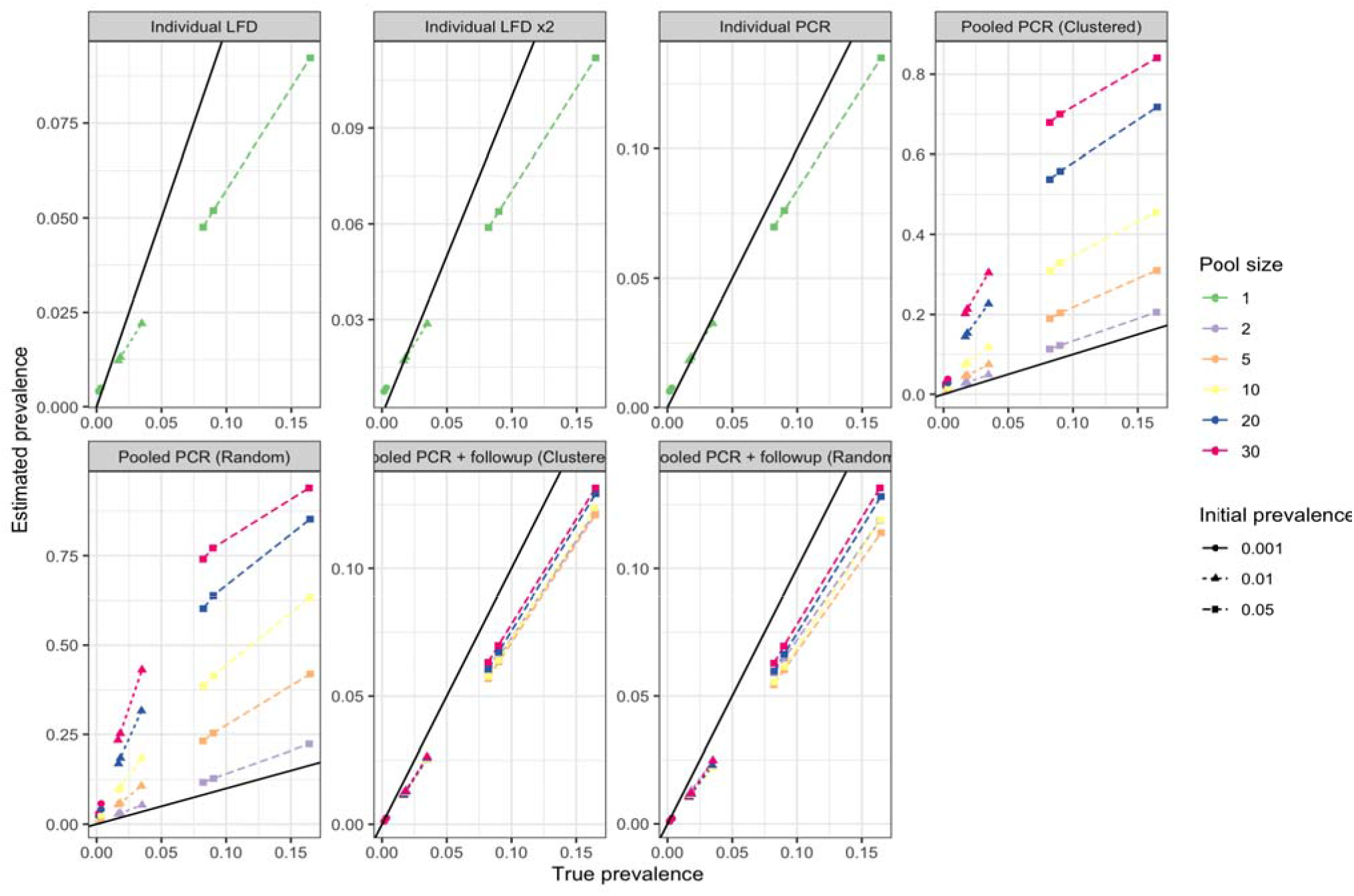
Comparison of prevalence estimates across testing strategies. Each plot represents a different testing strategy, the x-axes represent the true prevalence at the time of sampling, and the y axes represent the prevalence estimated by that testing strategy. For the pooled PCR strategy, it is assumed that everyone within the pool is positive.

## Discussion

A regular testing regimen across the student body may demand test pooling as the only viable strategy that financial and reagent resource limits permit. We present a framework which can be adapted to accommodate changes in estimates of viral load and RT-qPCR operating parameters, such as efficiency and sensitivity, as these estimates emerge and evolve. This study illustrates that there are trade-offs to be made in using this approach, which may require ethical arguments to inform decision making. In particular, reducing testing costs will incur a modest reduction in sensitivity, and some of the lowest cost pooling approaches have low specificity which could lead to large numbers of students self-isolating whilst not actually being infected. We note that the current pilot project for voluntary pooled testing in UK university students places students in pool sizes of up to 5 individuals in a household and follows up any positive pooled samples^11^. This strategy appears to be the most cost effective across scenarios in our simulations, including when compared against the LFD.

Pooling samples in a way that is informed by social contacts appears to be an effective strategy to improve pooling performance but does come at a cost of higher management and organisational cost at the levels of sample collection and laboratory systems. Clustered samples may however fall into the same pools without a management layer required, for example if samples from the same classroom or hall of residence are collected together. In these simulations we used living circle as a predictor of case clustering, and note that though it is an imperfect predictor it improves performance over random pooling. Any management scenario that is able to group individuals into test pools based on shared exposures (or shared lack of exposures) will likely improve pooling performance above the random case.

A key limitation of the costing analysis is that the logistics and time required to pool samples using maximal bin packing was not considered. It’s likely that the most realistic way to achieve pooling by living circle would be for samples to be submitted together and grouped within the lab according to when they are submitted. The true cost per PPV of pooling is therefore likely to be affected by the limitations of implementing pooling. Similarly, practical considerations within the laboratory infrastructure do place limitations on the complexity of the sample grouping strategies.

Per-individual follow-up testing within positive pools improves the specificity of test results but can become costly as prevalence increases. Outbreaks have been seen to be controlled within university settings (e.g. ^27^) which suggests that structural measures are effective and it is unlikely that prevalence will ever be particularly high at any given point^5^. Given that close contacts of positive cases are advised to self-isolate regardless of their test status, follow-up testing of positive pools may provide unactionable information when then pools are composed of social contacts. However, the value of testing the contacts of infected individuals has recently been shown in other simulations^28^, where it was suggested that testing of contacts is likely to identify substantially more true negatives than false negatives.

Changes in the ratio of cost of sampling compared to cost of PCR test will affect the difference in costs between pooling sizes, and if PCR test costs are reduced then the advantages of pooling will be limited. However, if reagent supply is limited, this may drive up the cost of PCR tests while also providing further incentive to pool samples.

Our simulations indicate that the LFD has sensitivity and specificity profiles comparable to those of higher order pooling strategies. Emerging evidence does appear to contradict this finding, in which it has been suggested that LFD has substantially worse sensitivity than RT-qPCR^29^. There are three ways in which our simulations could be over-estimating the sensitivity of LFD. First, if in practice LFD is routinely performed using self-administered swabs and RT-qPCR is performed by a trained technician then this will have an adverse consequence on the sensitivity of LFD. Second, if the viral distribution follows a different shape to that which we assume, in particular with a large mass in the range of 30 < *C*_*q*_ < 35, then this is the range in which LFD has been suggested to have a substantial drop off in sensitivity^13^. Third, the relationship between LFD sensitivity and RT-qPCR *C*_*q*_ values may be biased upwards in reference ^13,30^.

There are a number of limitations to this study. First, we do not have data on the true heterogeneity of viral load amongst infected students, and higher heterogeneity could have unpredictable effects on the efficacy of pooling by living circle. If individuals with higher viral load are more infectious then the pooling of close contacts with those individuals will potentially increase sensitivity. Second, we used a theoretical framework to infer the RT-qPCR sensitivity beyond 10x dilution, and therefore the most reliable pool size within these simulations is at 10x dilution. However, we note that the projected sensitivity of the test beyond 10x dilution is in line with sensitivity estimates from experiments published previously^31^. Third, we only used a simple 2-time-step model to simulate clustering. The intention behind this approach was to simply obtain different levels of clustering of cases, but the true extent of clustering is not known. We show that the improvements in performance of pooling by living circle is attenuated when cases are less clustered (**Supplementary figure 3**), and will tend towards the random pooling values. Additionally, if tests were performed regularly then one might expect that there will not be enough time for tertiary infections to occur before infected cases are discovered. Fourth, alternative viable approaches in addition to RT-qPCR and LFD such as reverse transcription loop-mediated isothermal amplification^32,33^ have not been modelled here.

In this study our simulation is predicated on the detection of infected individuals. However, for the purposes of controlling the spread of the disease whilst limiting unnecessary quarantines, those who have very low viral load are much more likely to be post-infectious or non-infectious than they are to be pre-infectious^34^. Therefore, approaches such as LFD or large pool sizes may not lead to substantially higher transmission and yet they could be substantially cheaper. On the comparison between LFD and pooling RT-qPCR, LFD has the advantage of being substantially faster, but operationally its sensitivity appears to be lower than initial laboratory testing indicated^35^. By contrast, pooling of samples in practice may in fact exceed the sensitivity performance that we assumed in these simulations^31^.

It is also of note that the new Variant of Concern 202012/01 (VOC), which is thought to have originated in the UK in late Summer or early Autumn 2020, and which has spread rapidly in the UK since then (even during the November lockdown), is estimated to be between 1.4 and 1.8 times more transmissible than the original (wild-type) SARS-CoV-2^36^. Early results suggest that this new variant is associated with significantly higher viral loads in the upper respiratory tract, than was the case with wild-type SARS-CoV-2^37^. If a higher viral load is maintained across the infection course, then all tests will gain in sensitivity, and adopting pooling approaches (or the LFD) will have a reduced adverse impact on sensitivity when compared against the single RT-qPCR test.

Overall, we show that RT-qPCR pooling strategies are likely to be improved if individuals can be pooled based on likelihood of joint infection, and this could be achieved by pooling within known living or contact circles. Though we used student living circles as a basis for the simulations, the results could extend to households in general. An adaptive strategy, whereby different pooling schemes are used depending on the estimated prevalence and R values (e.g. from local authority reporting), could be optimal.

## Data Availability

All simulations were conducted using code from https://github.com/explodecomputer/covid-uob-pooling.

## Acknowledgements

We are grateful to Adam Finn for comments on an early version of the model. GH is funded by the Wellcome Trust [208806/Z/17/Z]. HC and EBP would like to acknowledge support from the National Institute for Health Research (NIHR) Health Protection Research Unit (HPRU) in Behavioural Science and Evaluation at the University of Bristol. HC is additionally funded through an NIHR Career Development Fellowship [CDF-2018-11-ST2-015]. The views expressed are those of the author(s) and not necessarily those of the NIHR or the Department of Health and Social Care. CR is a member of the MRC Integrative Epidemiology Unit and receives support from 501 the MRC (MC_UU_00011/5) and the University of Bristol. EN and EBP are supported by the EPSRC (MR/V038613/1). ATh is supported by Wellcome (217509/Z/19/Z).

## Author contributions

GH, JGW and AT wrote the first draft of the manuscript. GH and JGW conducted the simulations. EBP and GH had the original idea for the manuscript. GH, JGW, ACT, AT, EN, DE, RK, CR, AF, LD, HC and EBP designed analyses, interpreted the results, and critically reviewed the manuscript.

