## Supplementary material for "Modelling pooling strategies for SARS-CoV-2 testing in a university setting": Suplpementary information

Supplementary materials: Modelling pooling strategies for SARS-CoV-2 RT-qPCR testing in a university setting

Hemani et al

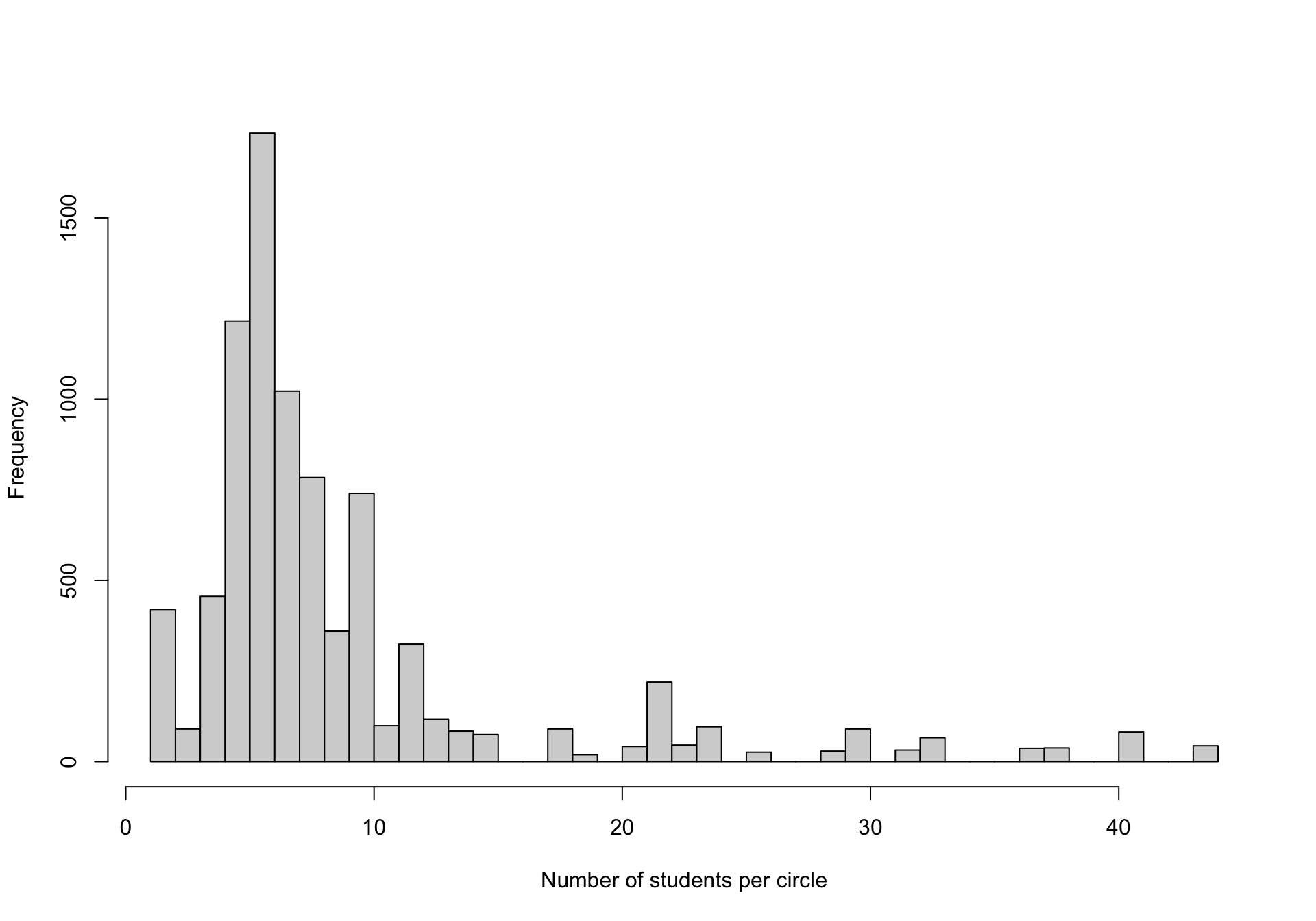
**Supplementary figure 1**: Number of students per living circle

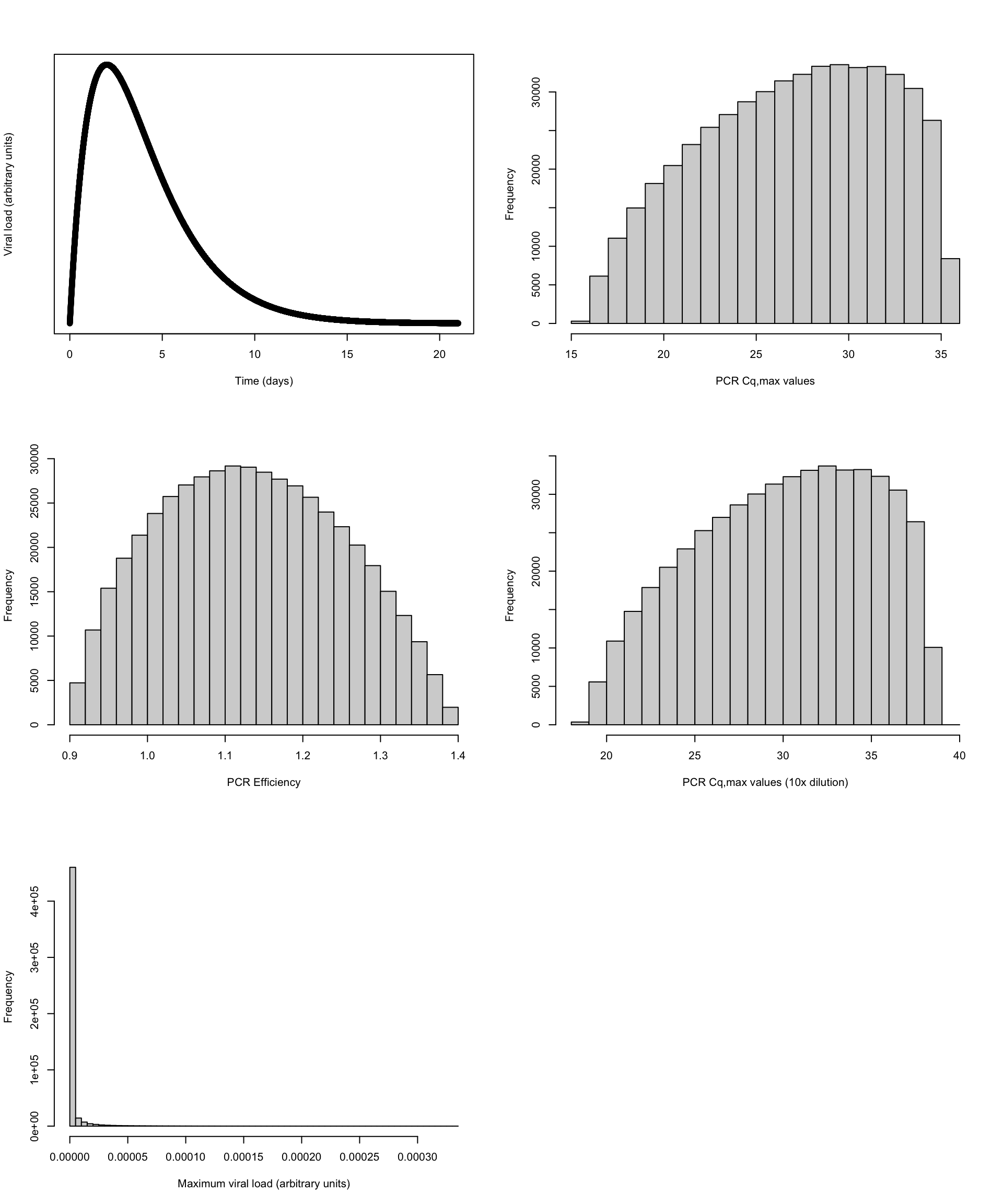

**Supplementary figure 2: Distributions relating to the RT-qPCR kinetics based on parameter values that are determined following optimisation.** Top left: Viral load during infection time course. Top right: distribution of $C_{q,max}$ values for undiluted positive samples. Middle left: Distribution of reaction efficiencies. Middle right: Distribution of $C_{q,max}$ values for positive samples diluted by 10x. Bottom left: Distribution of viral load amongst positive samples in the population. Y-axis represents frequencies for arbitrarily large samples.

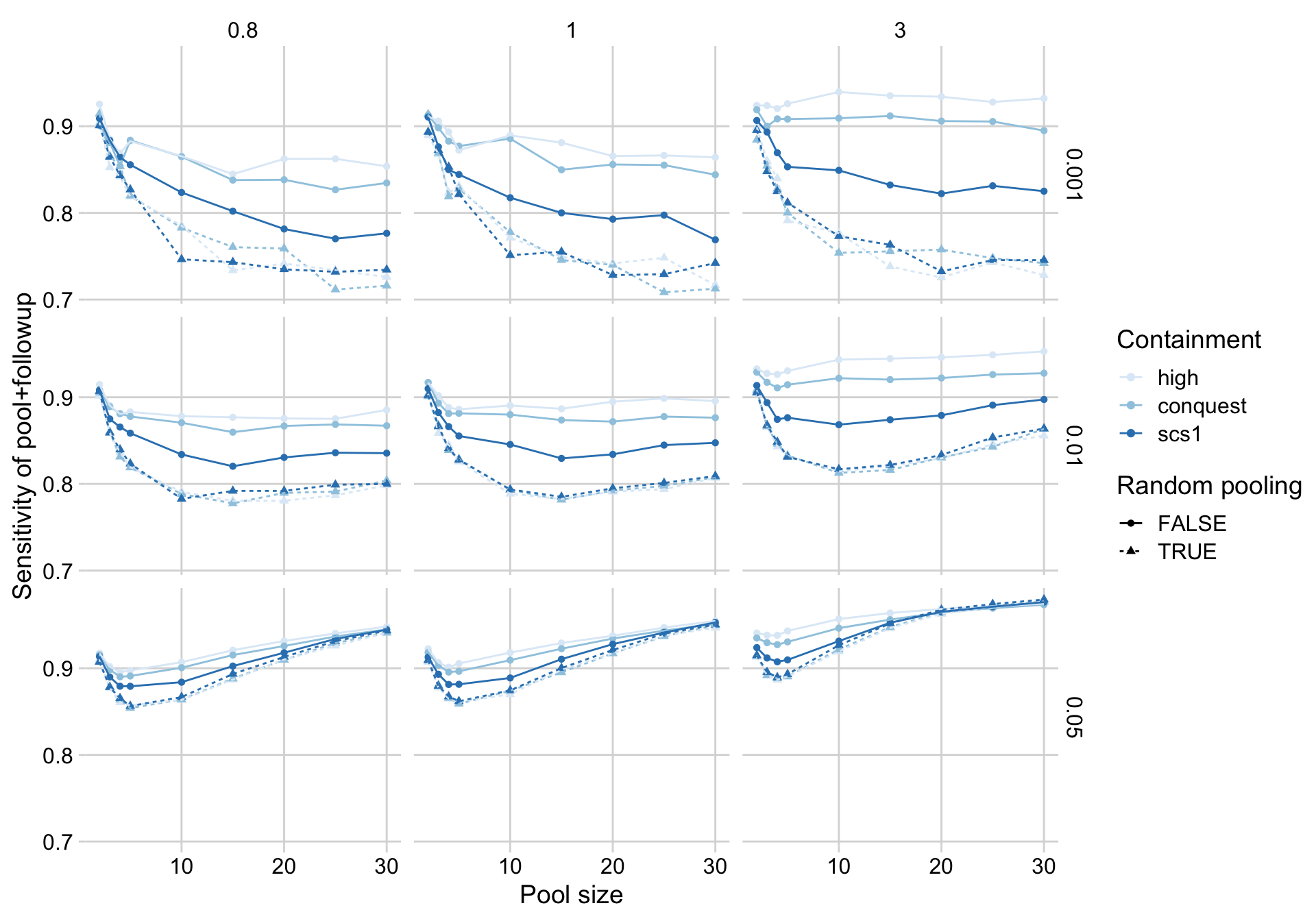

**Supplementary figure 3: Changes in sensitivity of pooling depending on the containment scenario modelled.** Throughout the text we use the CON-QUEST transmission event probabilities, and in this figure we compare the sensitivity of the pooling + follow up model from this model against two other models – a ‘high containment’ model where students are very unlikely to transmit outside of their living circle, and the ‘scs1’ model which is based on student mixing patterns surveyed outside of the pandemic context, and transmission within living circles is substantially reduced. All scenarios indicate that pooling based on living circles outperforms random pooling.

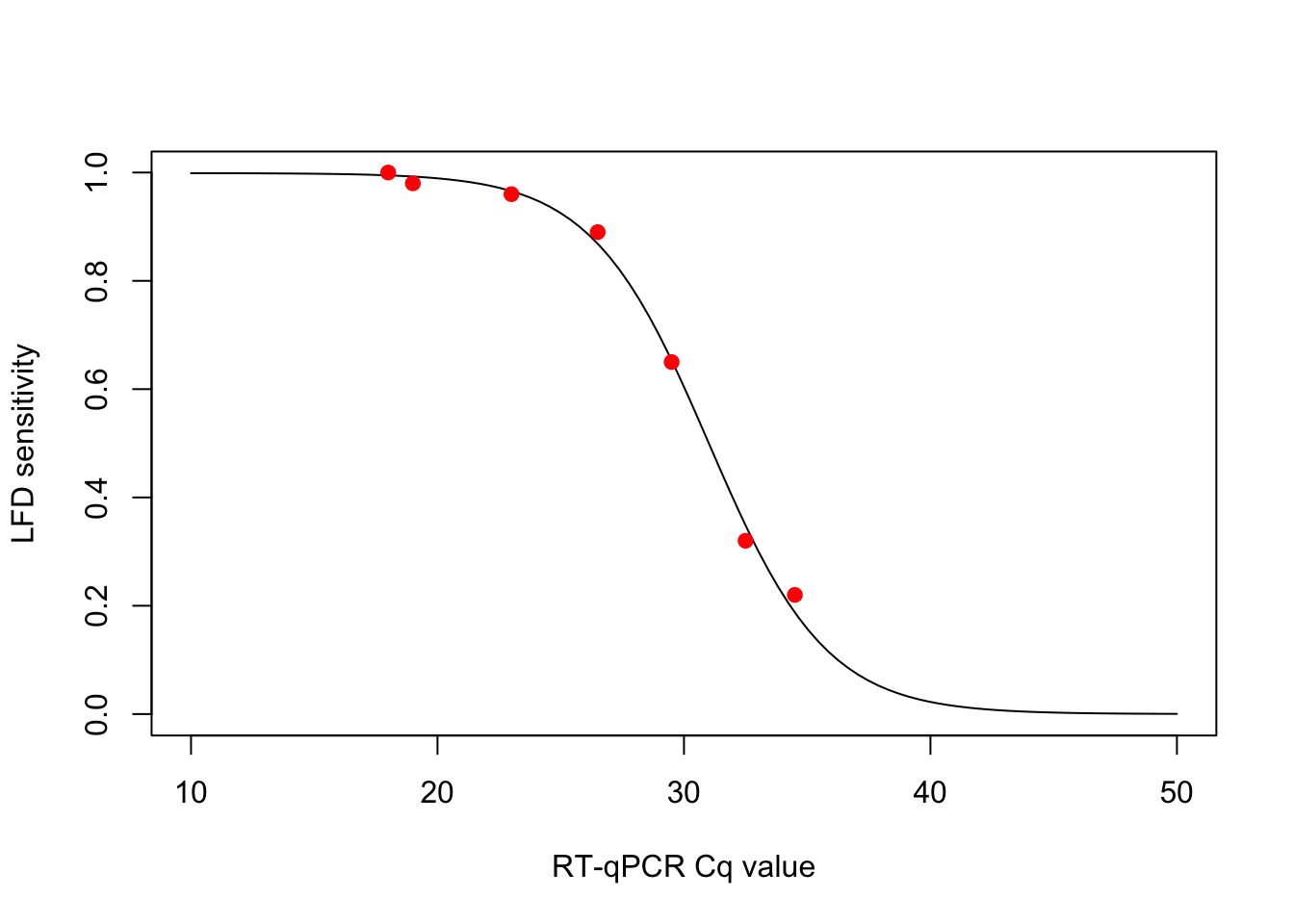

**Supplementary figure 4:** Relationship between RT-qPCR $C_{q}$ value and the LFD sensitivity. Red points are taken from figure 2 of the Joint PHE Porton Down & University of Oxford SARS-CoV-2 test development and validation cell rapid evaluation of the LFD. Sigmoid curve is an interpolation against these points.
